# Reverse Transcriptase Loop Mediated Isothermal Amplification (RT-LAMP) for COVID-19 Diagnosis: A Systematic Review and Meta-Analysis

**DOI:** 10.1101/2021.01.04.20248979

**Authors:** Anita Dominique Subali, Lowilius Wiyono

## Abstract

**Background:** Coronavirus Disease 2019 (COVID-19) has caused a severe outbreak and become a global public health priority. Rapid increment of infection number along with significant deaths have placed the virus as a serious threat to human health. Rapid, reliable, and simple diagnostic methods are critically essential for disease control. While Reverse Transcriptase Polymerase Chain Reaction (RT-PCR) is the current diagnostic gold standard, Reverse Transcriptase Loop-Mediated Isothermal Amplification (RT-LAMP) appears as a compelling alternative diagnostic test due to its more simplicity, shorter time to result, and lower cost. This study examined RT-LAMP application for rapid identification of SARS-CoV-2 infection compared to RT-PCR assay.

**Methods:** A systematic review and meta-analysis (2020) was conducted in 6 scientific databases following the PRISMA Guideline. Original published studies on human clinical samples in English were included. Articles evaluated sensitivity and specificity of RT-LAMP relative to RT-PCR were considered eligible. Quality assessment of bias and applicability was examined based on QUADAS-2.

**Results:** A total of 351 studies were found based on the keywords and search queries. 14 eligible case control studies fitted the respective criteria. Quality assessment using QUADAS-2 indicated low risk bias in all included studies. All case studies, comprises 2,112 samples, had the cumulative **sensitivity** of **95.5% (CI 97.5%=90.8-97.9%)** and cumulative **specificity** of **99.5% (CI 97.5%=97.7-99.9%)**.

**Conclusion:** RT-LAMP assay could be suggested as a reliable alternative COVID-19 diagnostic method with reduced cost and time compared to RT-PCR. RT-LAMP could potentially be utilized during the high-throughput and high-demand critical situations.

## INTRODUCTION

Recently emerging Novel Corona Virus Disease (COVID-19), the subsequent disease due to Severe Acute Respiratory Syndrome Coronavirus 2 (SARS-CoV-2) infection has created a skyrocketing outbreak and become a serious threat towards global public health emergency(1). As of 3 January 2021, the COVID-19 pandemic has resulted in a total of 83,322,449 confirmed cases, including 1,831,412 deaths according to the WHO COVID-19 report (2) SARS-CoV-2 currently known to be transmitted among humans by the means of respiratory droplets and aerosols produced from infected persons while sneezing, talking, and/or coughing (3). At the moment, effective antiviral drugs and vaccines for SARS-CoV-2 are still under research (4) The absence of suitable therapeutic agents necessitates prompt diagnosis with simple, rapid, reliable detection of SARS-CoV-2 infection (5).

Diagnosis of early infection is difficult since COVID-19 manifests with non-specific clinical symptoms, such as fever, cough, or shortness of breath, which overlap those found at common cold and influenza, even though some progressed to severe complication (1,3). In addition, patients with early SARS-CoV-2 infection can remain asymptomatic. Symptoms appear as early as 2 days or up to 2 weeks after exposure (4). Hence, the confirmation of SARS-CoV-2 infection relies on nucleic acid testing, which detects the viral RNA (6). Early, rapid, and accurate identification of SARS-CoV-2 infected patients, even before immune response occurred and for asymptomatic carrier, is crucial, not only to provide appropriate timely medical support for patients but also to take mitigation actions to impede further spread and control transmission (1,7,8).

The current gold standard for COVID-19 molecular diagnosis is based on real-time reverse transcription-polymerase chain reaction (RT-PCR), which detects SARS-CoV-2 RNA (6). RT-PCR produces outstanding analytical performances with highly sensitive and specific results (8). Nonetheless, RT-PCR based approaches for COVID-19 detection still suffer from several limitations, since these methods require highly skilled personnel and sophisticated equipment with poor availability (restricted to public health laboratories) (4). As a result, they are considered impractical, especially in the setting of remote areas and developing countries with limited resources (3). Yet, the fact that PCR-based methods are relatively time consuming with complicated processes limits their capacity to meet the demand in the active pandemic situation (3).

Rapidly growing number of incidences urges substitution with an equally reliable molecular detection method to facilitate detection of SARS-CoV-2 infection (1,3). In order to answer this challenge, a test kit which offers shorter turn-around time and is simple to operate would be highly desirable. The test should be able to identify virus infected patients, even at an early stage. Ideally, the diagnostic test is mobile, reduces the need of any complicated instrument, and the result can be interpreted by the naked eye. Therefore, it is feasible to be used at public facilities, particularly at health centers in rural areas (9).

Reverse Transcriptase Loop-mediated isothermal amplification (RT-LAMP) is a rapid one-step DNA amplification technique (7). This method has been applied to numerous pathogen detection, such as virus, bacteria, and malaria (9). RT-LAMP is regarded as a promising point-of-care test due to several advantages: high sensitivity and specificity, rapid reaction, and less dependent on sophisticated equipment (5). The LAMP reaction takes place at constant temperature at 60 to 65°C within less than an hour (8). Since the DNA amplification process occurs at one constant temperature, it requires simple equipment and eliminates the needs of thermal cycle as of PCR (6). The single reaction of RT-LAMP significantly shortens the reaction time which bypasses the DNA purification step. Hence, a rapid detection of SARS-CoV-2 can be achieved (9). RT-LAMP results can be detected by visual turbidity or fluorescence in real time (7). Therefore, the result can be visualized by naked eye and interpreted by any-person within a short turn-around time(5). Last but not least, the simple procedure allowed this skill to be easily mastered by junior laboratory technologists or healthcare workers with short training (8).

Regarding these facts, a review was conducted in order to examine diagnostic performance of RT-LAMP compared to RT-PCR, the current diagnosis gold standard of COVID-19 diagnosis. By the time of the review was performed, authors had not found other similar studies analyzed RT-LAMP for COVID-19. The review results were expected to provide credible evidence of the use of the proposed diagnostic tool, RT-LAMP, as a potential alternative to answer the current issue.

## MATERIAL & METHODS

This systematic review and meta-analysis followed the Preferred Reporting Items for Systematic Review and Meta-Analyses (PRISMA) guidelines (10) to properly search for relevant studies or literatures used in this review

### Search strategy

Two independent investigators performed thorough literature searches in a blinded fashion. Discrepancies were resolved by the discussion of both investigators. The literature search was done through seven scientific databases, namely Pubmed, ScienceDirect, Scopus, Proquest, Wiley, Scopus, and EBSCohost for studies published up to October 12th 2020. The literature search strategies were developed using medical subject headings (MeSH) in scientific databases with text words related to COVID-19 diagnosis, RT-PCR, and RT-LAMP. The used keywords in each database are listed on **Table 1**. We also do a manual hand-searching reference list from the included studies. Subsequently, the retrieved results were deduplicated and screened against the pre-specified eligibility criteria.

**Table 1.** Keywords or queries used in each database for the literature search process

| Database | Queries |
| --- | --- |
| PubMed | ((PCR [MeSH] **OR** “Reverse Transcriptase Polymerase Chain Reaction” [Title/Abstract]) **AND** (RT-LAMP [Title/Abstract] **OR** “Reverse Transcriptase Loop Mediated Isothermal Amplification” [Title/Abstract])) **AND** (SARS-COV-2 [MeSH] **OR** COVID-19 [MeSH]) |
| Scopus | ((PCR **OR** “Reverse Transcriptase Polymerase Chain Reaction”) **AND** (RT-LAMP **OR** “Reverse Transcriptase Loop Mediated Isothermal Amplification”)) **AND** (SARS-COV-2 **OR** COVID-19) |
| ProQuest | ((PCR **OR** “Reverse Transcriptase Polymerase Chain Reaction”) **AND** (RT-LAMP **OR** “Reverse Transcriptase Loop Mediated Isothermal Amplification”)) **AND** (SARS-COV-2 **OR** COVID-19) |
| ScienceDirect | ((PCR **OR** “Reverse Transcriptase Polymerase Chain Reaction”) **AND** (RT-LAMP **OR** “Reverse Transcriptase Loop Mediated Isothermal Amplification”)) **AND** (SARS-COV-2 **OR** COVID-19) |
| EBSCOHost | ((PCR **OR** “Reverse Transcriptase Polymerase Chain Reaction”) **AND** (RT-LAMP **OR** “Reverse Transcriptase Loop Mediated Isothermal Amplification”)) **AND** (SARS-COV-2 **OR** COVID-19) |
| Wiley | ((PCR **OR** “Reverse Transcriptase Polymerase Chain Reaction”) **AND** (RT-LAMP **OR** “Reverse Transcriptase Loop Mediated Isothermal Amplification”)) **AND** (SARS-COV-2 **OR** COVID-19) |

### Study Eligibility Criteria

Inclusion criteria was set to filter primary studies investigating the diagnostic study of RT-LAMP in COVID-19 diagnosis using PICO (Patient, Intervention, Control, Outcome) criteria as summarized in **Table 2**. We included studies consisting of prospective and retrospective, cross-sectional, and cohort published studies from human clinical samples of COVID-19 suspected patients, which evaluated sensitivity and specificity of RT-LAMP for COVID-19 diagnosis in comparison to RT-PCR. Meanwhile, studies were also excluded if any of the following criteria were met: (1) Review articles, case series, or letter to editors; (2) in-vitro studies without clinical samples; (3) irretrievable full-text articles; and (4) non-English articles.

**Table 2.** PICO criteria used, consists of four parameters: patient, intervention, control, and objective criteria

| Parameter of PICO | Inclusion criteria |
| --- | --- |
| Patient | Suspected or confirmed COVID-19 patients |
| Intervention (Index test) | RT-LAMP Kit |
| Control (Reference test) | RT-PCR |
| Objective | Sensitivity & Specificity (Diagnostic value) |

### Study Selection

After the previous search in proposed databases had been completed, the search results were stored in Microsoft Excel Spreadsheet (Microsoft Corp, Redmond, WA, USA). Authors manually checked and removed duplicate articles. The spreadsheet was also utilized to manage the data. The authors independently reviewed the literature search. Primarily, a screening on titles and abstracts of the selected articles was conducted to exclude studies according to the exclusion criteria. The reviewers mentioned the underlying reason to exclude any items in the spreadsheet. Studies were included for the following step if there was any uncertainty. Subsequently, the authors read the full text in order to exclude studies which did not meet inclusion criteria. The selected studies were then validated by conducting a meeting among the reviewers to select which article was considered eligible for review. A data extraction form was built to compile data from the included studies.

### Data Extraction and Quality Assessment

Two reviewers (ADS, LW) independently collected the data. Data were justified and discussed by both authors to ensure completeness and plausibility before being synthesized. Variables were recorded such as: authors, year published, location of study, study design, sample size, study sample/population, index test (RT-LAMP), reference test (RT-PCR), clinical setting (inpatient vs outpatient), and level of evidence. The main outcomes for this review were the diagnostic values of RT-LAMP, consisting of the true positive (TP), true negative (TN), false positive (FP), and false negative (FN) of all studies, consequently their sensitivity and specificity value.

The included studies were then assessed for their methodological quality to reduce systematic biases and inferential errors of the data extracted. Two reviewers (ADS, LW) independently assessed risk of bias of the included studies using Quality Assessment of Diagnostic Accuracy Studies (QUADAS-2) (11). There are four areas of bias assessed in QUADAS-2 (11): patient selection, index test, reference standards, and flow/timing. All the studies were subsequently judged to be yielding low, unclear, and high risk of bias. The authors also assessed the concerns of applicability from three areas: patient selection, index test, and reference standards, which subsequently judged to be yielding low, unclear, and high concerns of applicability. The summary of risk of bias and graph were generated in Microsoft Excel Spreadsheet.

### Statistical Analysis

Quantitative analysis was performed using the outcome of studies extracted from the included studies. Values of test accuracy were compared with RT-PCR methods, including sensitivity, specificity, true positive (TP), false negative (FN), false positive (FP), and true negative (TN) – using data extracted from sources or calculated from the available data. RT-PCR was considered as a reference test and comparator to the index test, RT-LAMP. Results of individual studies were graphically presented on forest plot, as well as Diagnostic Value and random effects curve. Meta-analysis was conducted using MetaDTA software (University of Leicester, Leicester, England).

## RESULTS

### Study selection and characteristics

The literature search process is summarized in **Figure 1**, using PRISMA Guideline (10) flowchart. The initial search yielded 364 records, of which 16 articles were deduplicated and 332 articles were excluded following the title and abstract screening. A total of 16 articles were then properly assessed on full-text assessments, which resulted in two articles being excluded due to inappropriate study design.

**Figure 1.**
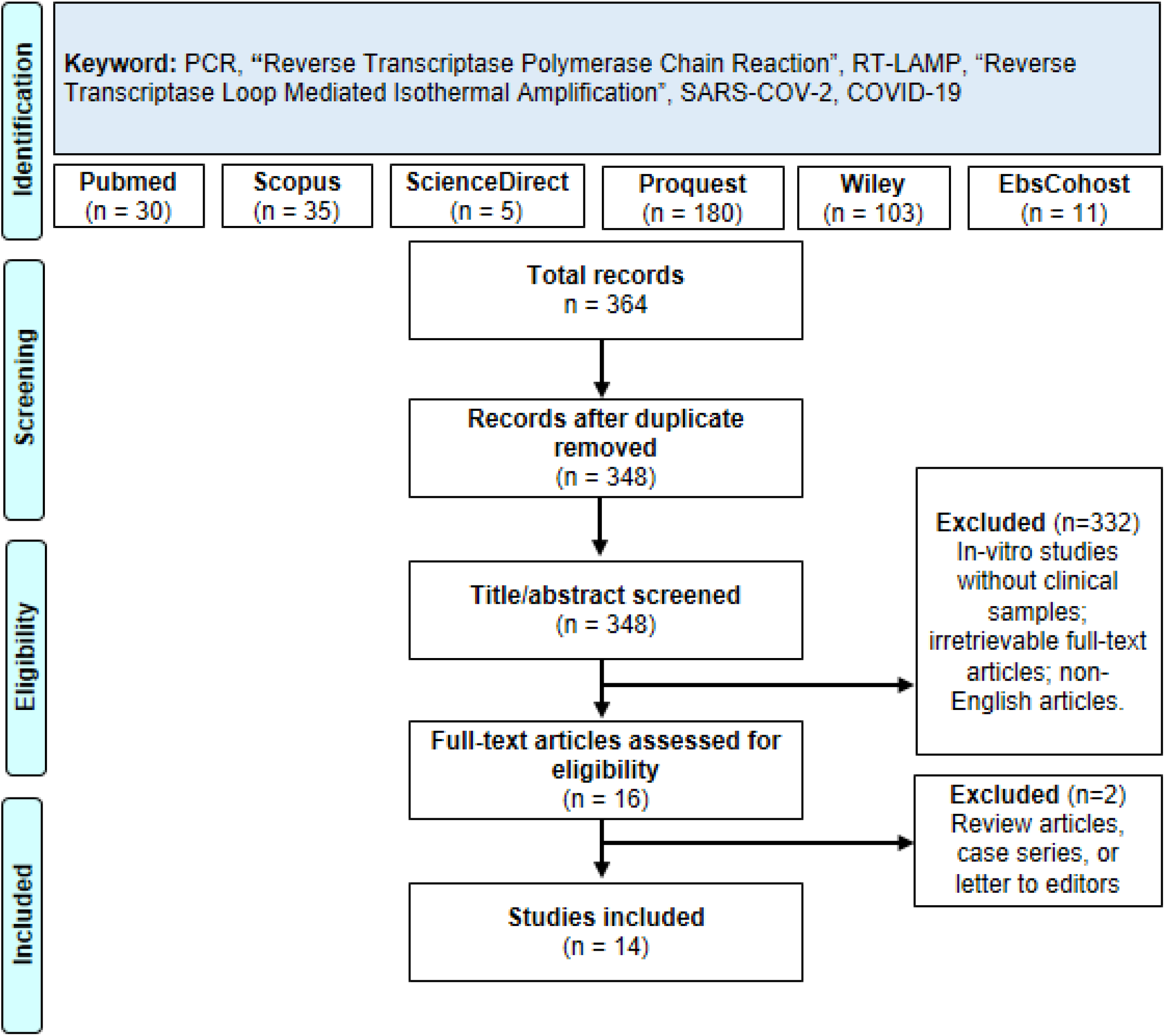
Literature search strategy based on PRISMA Guideline

Consequently, a total of 14 articles were included, comprising 2,112 patients or samples, consisting of either suspected or confirmed COVID-19 patients. Patients’ characteristics are not mentioned in most studies therefore the authors didn’t extract samples’ characteristics. The included studies consist of quite a variety of samples, of which there are five studies from China; two studies from Germany (12,13); and one study each from Hongkong (8), China (6), Japan (14), Malaysia (15), South Korea (7), Canada (16), Australia (17), and USA (4). As for the index test, there are seven studies using RT-LAMP Kit (1,4,7–9,14,15,18), while other studies used Warmstart RT-LAMP Assay Kit (n=2) (5,16), Variplex RT-LAMP Assay Kit (n=1) (12), Loopamp RNA Amplification Kit (n=1) (6), RT-LAMP Mastermix kit (n=1) (17), and specific colorimetric and fluorescence RT-LAMP Kit (n=1) (13). As for the reference test, most studies used the qRT-PCR Kit with two studies specified the used kit (BGI qRT-PCR Kit (19) and NPMA RT-PCR Kit (18)), while several studies used LightMix E-Gene RT-PCR Kit (n=3) (12,13,17). Most studies used the cross-sectional study design (n=13), with one exception using prospective cohorts in its study(1). The included studies’ characteristic is summarized in **Table 3**.

**Table 3.** Included studies characteristics

| Studies,<br>year | Studies characteristics |  |  |  |  |  |  |  |
| --- | --- | --- | --- | --- | --- | --- | --- | --- |
|  | Location | Design | Sample size | Population/ Sample | Index Test | Reference Test | Clinical settings | Level of evidence* |
| Mohon *et al*, 2020 | Calgary, Canada | Cross-sectional | 124 | COVID-19 patients/ Archived nasopharyngeal swab | Warmstart RT-LAMP assay | RT-PCR | N/A | 1C |
| Rodel *et al*, 2020 | Penzberg, Germany | Cross-sectional | 137 | COVID-19 patients/ Archived nasopharyngeal swab | Variplex RT-LAMP assay | LightMix E-Gene RT-PCR | N/A | 1C |
| Chow *et al*, 2020 | Hongkong | Cross-sectional | 362 | COVID-19 and other viral infection/Nasopharyngeal swab | COVID-19 RT-LAMP assay | qRT-PCR | Inpatient | 1C |
| Kitagawa *et al*, 2020 | Saitama, Japan | Cross-sectional | 76 | Suspected COVID-19 patient/ N/A | RT-LAMP Kit | qRT-PCR | Outpatient | 1C |
| Hu *et al*, 2020 | Guangdong, China | Cohort | 481 | Suspected COVID-19 patient/ N/A | RT-LAMP Kit | qRT-PCR | Inpatient | 1B |
| Jiang *et al*, 2020 | Wenzhou, China | Cross-sectional | 260 | COVID-19 and other viral infection/Nasopharyngeal swab | RT-LAMP Kit | NPMA qRT-PCR | N/A | 1C |
| Yan *et al*, 2020 | Beijing, China | Cross-sectional | 130 | COVID-19 and pneumonia patients/Nasopharyngeal swab | Loopamp RNA amplification kit (RT-LAMP) | BGI RT-PCR Kit | N/A | 1C |
| Lamb *et al*, 2020 | Belmont, USA | Cross-sectional | 20 | N/A / Serum, urine, oropharyngeal, nasopharyngeal swab | RT-LAMP Kit | qRT-PCR | Outpatient | 1C |
| Lu *et al*, 2020 | Nantong, China | Cross-sectional | 56 | Suspected COVID-19 patients/Nasopharyngeal swab | Warmstart RT-LAMP Assay | RT-qPCR kit | Inpatient | 1C |
| Baek *et al*, 2020 | Seoul, South Korea | Cross-sectional | 154 | COVID-19 and other viral infection/ Nasopharyngeal swab | RT-LAMP kit | qRT-PCR | N/A | 1C |
| Lee *et al*, 2020 | Melbourne , Australia | Cross-sectional | 157 | N/A / Nasopharyngeal swabs | RT-LAMP Mastermix | E-Gene RT-qPCR | N/A | 1C |
| Klein *et al*, 2020 | Heidelberg, Germany | Cross-sectional | 77 | Random eligible sample/ Nasopharyngeal swab | Colorimetric and Fluorescence RT-LAMP | E-Gene RT-qPCR | N/A | 1C |
| Lau *et al*, 2020 | Sungai Buloh, Malaysia | Cross-sectional | 89 | COVID-19 patients/ Archived nasopharyngeal swab | RT-LAMP Kit | qRT-PCR | N/A | 1C |
| Huang *et al*, 2020 | Ningbo, China | Cross-sectional | 16 | COVID-19 and other viral infection/Nasopharyngeal swab | RT-LAMP Kit | qRT-PCR | Outpatient | 1C |

### Risk of Bias Assessment

The risk bias assessment using QUADAS-2 (11) revealed most studies yielded low and unclear risk of bias. On the patient selection aspects, 10 out of 14 studies had unclear risk of bias with 4 out of 14 showed low risk of bias. This result could be explained as most studies not explicitly declare their study design. While on the reference test aspect, there are 7 out of 14 studies with unclear risk of bias with another seven studies having a low risk of bias. The unclear risk of bias was decided due to the unclear statement of each study on the blinded fashion of data analysis on reference tests without knowing index test results. On index test assessment, 5 out of 14 studies had unclear risk of bias because four studies did not mention the blinded fashion of data extraction and one study had no pre-specified threshold on index test. On the flow and timing aspect, 3 out of 14 studies yielded unclear risk of bias due to unclear statements on all three studies on interval time between reference test and index test. All studies not mentioned above yielded low risk of bias.

Our review question did not focus on any particular patient demographics, and all of the included studies did not exclude patients based on their demographics characteristic, therefore, there was no concern of applicability for the patient selection. The index test, which is RT-LAMP was also not specifically mentioned in our review question, and therefore all RT-LAMP kit is applicable in our review. The reference standard tests in nearly all included studies also used qRT-PCR, as the current gold standard tests for COVID-19 diagnosis, and thus these studies were determined to have low concern of applicability in both index test and standard test. Therefore, all included studies yield low concern of applicability in all aspects. Summary of QUADAS-2 assessment could be found in **Figure 2**.

**Figure 2.**
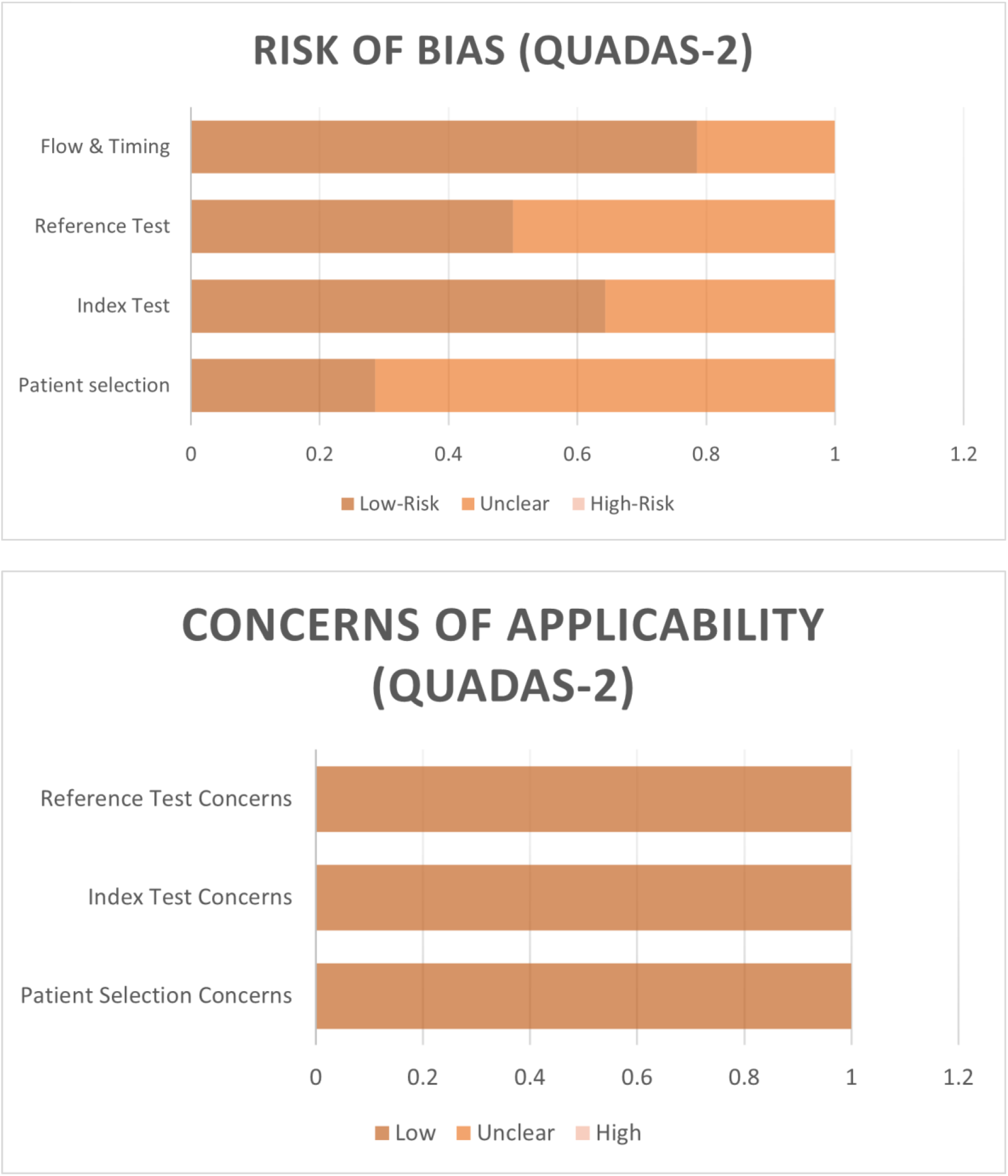
QUADAS-2 assessment on risk of bias and concerns of applicability

### Outcome of Studies

The summary of the outcome of each included study is listed in **Table 4**. Study outcomes, consisting of TP, FN, FP, and TN values were listed and the sensitivity and specificity value of each study were then calculated. Sensitivity of RT-LAMP is found to range from 75 to 100% while its specificity is found to range from 90 to 100%. All outcomes of each study are extracted from the cumulative outcome if the mentioned study used several subgroups for their analysis. Out of all 14 studies, 10 studies have a sensitivity value of more than 90%, with 4 of them having a sensitivity value of 100%. There are three studies with sensitivity values of more than 80%, and only one study with sensitivity below 80%. While for its specificity value, all studies showed a high number of specificity with 11 studies showing 100% specificity value, while other three studies show a specificity value of more than 98%.

**Table 4.**
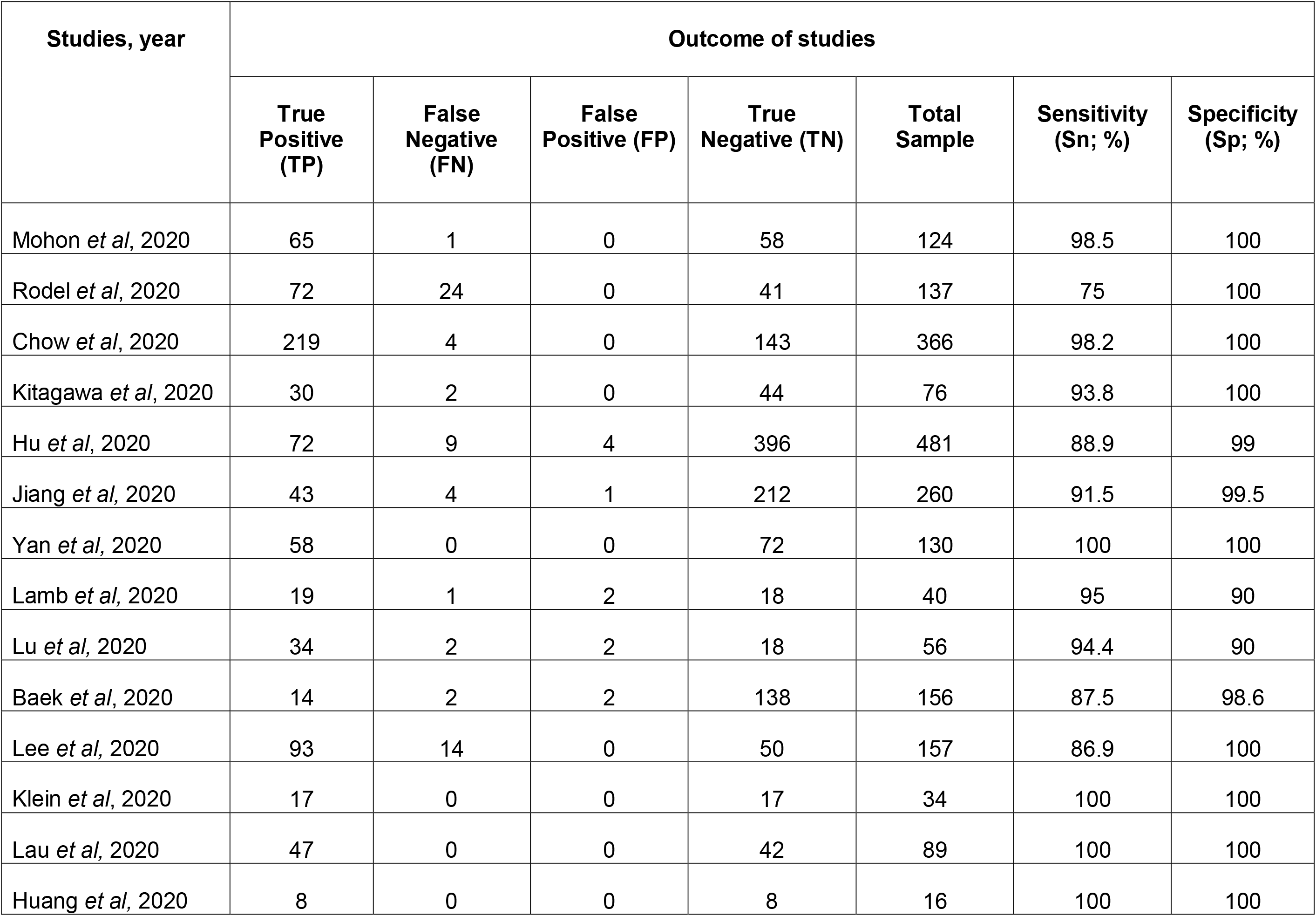
Outcome of studies

| Studies, year | Outcome of studies |  |  |  |  |  |  |
| --- | --- | --- | --- | --- | --- | --- | --- |
|  | True Positive (TP) | False Negative (FN) | False Positive (FP) | True Negative (TN) | Total Sample | Sensitivity (Sn; %) | Specificity (Sp; %) |
| Mohon *et al*, 2020 | 65 | 1 | 0 | 58 | 124 | 98.5 | 100 |
| Rodel *et al*, 2020 | 72 | 24 | 0 | 41 | 137 | 75 | 100 |
| Chow *et al*, 2020 | 219 | 4 | 0 | 143 | 366 | 98.2 | 100 |
| Kitagawa *et al*, 2020 | 30 | 2 | 0 | 44 | 76 | 93.8 | 100 |
| Hu *et al*, 2020 | 72 | 9 | 4 | 396 | 481 | 88.9 | 99 |
| Jiang *et al*, 2020 | 43 | 4 | 1 | 212 | 260 | 91.5 | 99.5 |
| Yan *et al*, 2020 | 58 | 0 | 0 | 72 | 130 | 100 | 100 |
| Lamb *et al*, 2020 | 19 | 1 | 2 | 18 | 40 | 95 | 90 |
| Lu *et al*, 2020 | 34 | 2 | 2 | 18 | 56 | 94.4 | 90 |
| Baek *et al*, 2020 | 14 | 2 | 2 | 138 | 156 | 87.5 | 98.6 |
| Lee *et al*, 2020 | 93 | 14 | 0 | 50 | 157 | 86.9 | 100 |
| Klein *et al*, 2020 | 17 | 0 | 0 | 17 | 34 | 100 | 100 |
| Lau *et al*, 2020 | 47 | 0 | 0 | 42 | 89 | 100 | 100 |
| Huang *et al*, 2020 | 8 | 0 | 0 | 8 | 16 | 100 | 100 |

Both sensitivity value and specificity value were then shown in the forest plot to picture the outcomes of all studies. The forest plot is pictured in **Figure 3**. As shown in the forest plot, each of the studies’ sensitivity and specificity value is shown with a confidence interval (CI) of 95%.

**Figure 3.**
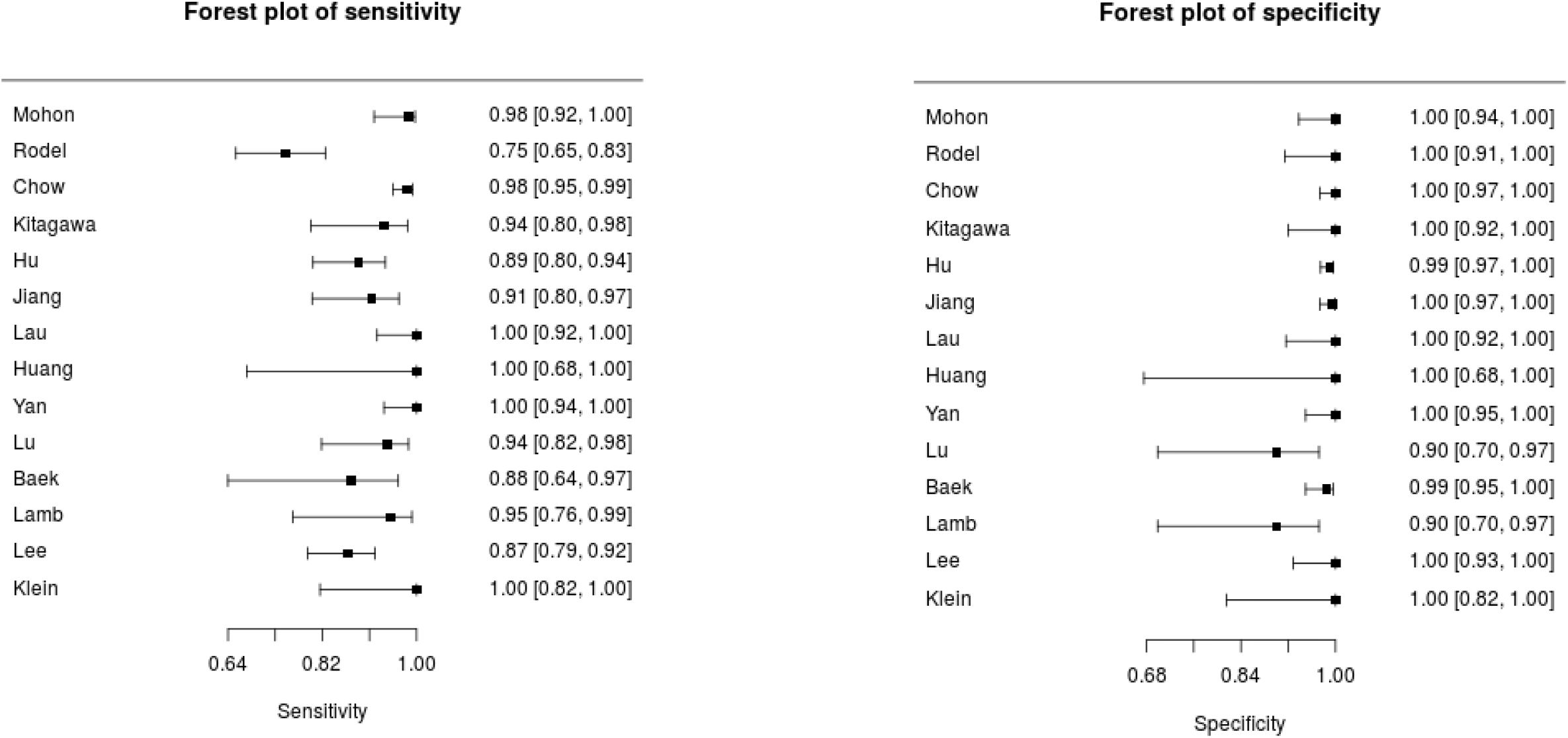
Forest plot of sensitivity and specificity of RT-LAMP on included studies

### Meta-Analysis of Sensitivity and Specificity

We performed a pool analysis on 14 included studies which used RT-LAMP as the index test and RT-PCR as the reference test. On the pooled analysis, we only estimate the pooled sensitivity and specificity for all studies and report the data in a SROC curve, as seen on **Figure 4**. Pooled analysis result is summarized in **Table 5**. Pooled sensitivity and specificity are at 99.5% (CI 97.5%: 90.8%-97.9%) and 95.5% (CI 97.5%: 97.7%-99.9%) respectively, indicating an overall good performance by RT-LAMP as a diagnosis test so far. The false positive rate is also considered low at 0.5% (CI 97.5%: 0.1%-0.23%)

**Table 5.** Pooled analysis outcome of meta-analysis on 14 studies, summarized in this table with sensitivity, specificity, false positive rate, and logit of sensitivity and specificity

| Pooled Analysis Outcome (n=14) |  |  |  |
| --- | --- | --- | --- |
| Parameter | Estimate | 2.5% CI | 97.5% CI |
| Sensitivity | 0.955 | 0.908 | 0.979 |
| Specificity | 0.995 | 0.977 | 0.999 |
| False Positive Rate | 0.005 | 0.001 | 0.023 |
| logit(sensitivity) | 3.053 | 2.286 | 3.819 |
| logit(specificity) | 5.26 | 3.738 | 6.782 |

**Figure 4.**
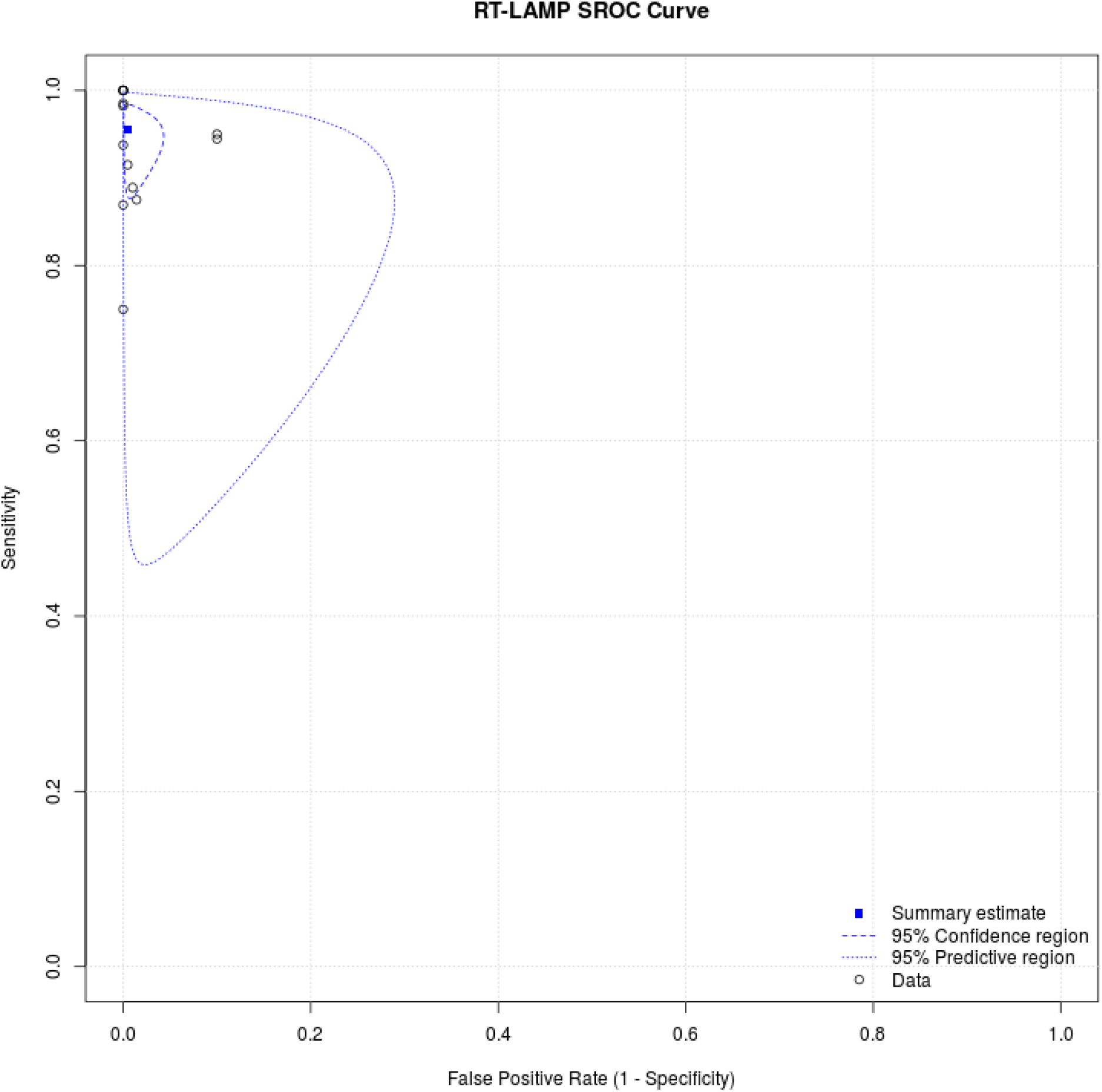
Summary Receiver Operating Characteristics (SROC) Curve on pooled analysis of RT-LAMP compared with RT-PCR

## DISCUSSION

Based on authors the latest search, this is the first systematic review and meta-analysis examining the performance and diagnostic value of RT-LAMP as a diagnostic test for COVID-19. The majority of included studies used RT-LAMP as the index test with RT-PCR as the reference test the sensitivity of at least 90%, with four studies reporting 100% sensitivity and specificity. While there are three studies with sensitivity values of <90% with one study reporting 75% sensitivity, we can safely assume that all studies reported a high diagnostic value of RT-LAMP on the diagnosis of COVID-19. At the time of this writing, the only published meta-analysis consisting of RT-LAMP for diagnosis was in the recent article by (20) which primarily investigated the performance of isothermal nucleic acid tests for human coronaviruses. Subsoontorn, *et al* (20) used 81 articles for the systematic review and 26 articles for subgroup analysis in the meta-analysis. The study (20) reported a pooled sensitivity at 0.94 (CI 95%: 0.90-0.96) for purified RNA from samples and sensitivity at 0.78 (CI 95%: 0.65-0.87) for crude samples. Our review shares the same result in performance of RT-LAMP with its high sensitivity value. However, our review does not analyze the difference of samples used in our analysis.

Among all studies, QUADAS-2 assessment has revealed a low or unclear risk of bias and concerns of applicability, with Kitagawa, *et al* (14) showed high quality study with low risk of bias and low concerns of applicability. However as most studies yielding low risk of bias, all studies are considered suitable to be included in the review and pooled analysis as there is no indication of high risk of bias or inapplicable study result.

While predominantly similar, there are some notable studies that evaluate the performance of RT-LAMP in various aspects. Rodel *et al* (12) performed a diagnostic test to assess RT-LAMP in various respiratory samples, compared with RT-PCR, and the combination of RT-LAMP and RT-PCR. Rodel (12) reported a combination of RT-PCR and RT-LAMP having sensitivity value at 92-100%. While the other 13 studies mention the diagnostic value of RT-LAMP kit alone compared with RT-PCR. Most studies used nasopharyngeal swab that has been obtained prior to the study, while Lamb, *et al* (4) used various samples, from respiratory samples to urine, however, the authors choose nasopharyngeal samples for the data extraction to provide similarity to other studies.

RT-LAMP is considered as a new and novel approach in diagnostic tests due to their rapid and low-cost kit Sahoo, *et al* (21), as mentioned by Ganguli, *et al* (22). It has been used for detecting both DNA and RNA viral pathogens, such as such as foot-and-mouth disease (23), human immunodeficiency virus (HIV), Japanese Encephalitis virus, Chikunguya virus, human papillomavirus, Denguevirus, West Nile virus, and Mumps virus (3). Furthermore, RT-LAMP has been reported effective for detecting other coronavirus, such as severe acute respiratory syndrome coronavirus (SARS-CoV) (24) and the Middle East respiratory syndrome coronavirus (MERS-CoV) (25).The most recent of the potential of using RT-LAMP as an emerging diagnostic test for COVID-19 was written in a mini-review by Thompson, *et al*. (26) and Kashir, *et al* (27). The lack of facilities in accommodating RT-PCR has given birth to cheaper and affordable diagnostic kits for COVID-19. Rodel *et al* (12)also mentioned a faster cycle of RT-LAMP, as there is no need for the thermal cycling process. However, all included studies have the same view on the potential of RT-LAMP.

Our study identified all relevant peer-reviewed studies published for deriving objective conclusions on the potential of RT-LAMP for the diagnosis of COVID-19. The authors have adhered this study to the standard guideline by PRISMA to ensure objective review and low risk of bias. However, this review has several limitations. The authors have seen that this review dominantly used RT-LAMP diagnostic value data on each study by extracting the cumulative data on all studies. Therefore, there was no extraction on subgroup analysis which has been used in some studies. In our search strategy, the authors used several databases to find peer-reviewed articles which are relevant to RT-LAMP diagnostic value on COVID-19. Systematic review and meta-analysis using preprints and ongoing clinical trials could be performed to gather more data for the systematic review, while keeping in mind to properly assess study quality. Subgroup analysis, particularly on types of RT-LAMP used in trials could also be used

## CONCLUSION

In conclusion, this systematic review and meta-analysis revealed the performance of RT-LAMP in diagnosing COVID-19 as compared with RT-PCR, its current gold standard diagnosis tools. A systematic review on 14 studies has shown a comparably high diagnostic value of RT-LAMP as seen in their sensitivity and specificity value. The pooled analysis on all included studies has revealed sensitivity value at 99.5% (CI 95%: 90.8%-97.9%) and specificity value at 95.5% (CI 95%: 97.7%-99.9%) respectively, thus the authors concluded high potential or performance of RT-LAMP in the diagnosis of COVID-19. The authors have declared several limitations, including only used peer-reviewed studies and no subgroup analysis. However, this leaves more improvement for further studies and reviews on the same respective topics.

## Data Availability

All data is available in the manuscript

## ACKNOWLEDGEMENTS

We would like to thank Jeremy Rafael Tandaju and Brenda Cristie Edina (Faculty of Medicine Universitas Indonesia, Jakarta, Indonesia) for the methodological and statistical advice.

## REFERENCES

1. Hu X, Deng Q, Li J, Chen J, Wang Z, Zhang X, et al. Development and Clinical Application of a Rapid and Sensitive Loop-Mediated Isothermal Amplification Test for SARS-CoV-2 Infection. mSphere [Internet]. 2020 Aug 26 [cited 2020 Oct 11];5(4). Available from: /pmc/articles/PMC7449630/?report=abstract

2. World Health Organization. WHO Coronavirus Disease (COVID-19) Dashboard WHO Coronavirus Disease (COVID-19) Dashboard [Internet]. 2021 [cited 2021 Jan 4]. Available from: https://covid19.who.int/

3. Augustine R, Hasan A, Das S, Ahmed R, Mori Y, Notomi T, et al. Loop-mediated isothermal amplification (Lamp): A rapid, sensitive, specific, and cost-effective point-of-care test for coronaviruses in the context of covid-19 pandemic [Internet]. Vol. 9, Biology. MDPI AG; 2020 [cited 2020 Oct 11]. p. 1–17. Available from: /pmc/articles/PMC7464797/?report=abstract

4. Lamb LE, Bartolone SN, Ward E, Chancellor MB. Rapid detection of novel coronavirus/Severe Acute Respiratory Syndrome Coronavirus 2 (SARS-CoV-2) by reverse transcription-loop-mediated isothermal amplification. PLoS One [Internet]. 2020 Jun 1 [cited 2020 Oct 11];15(6). Available from: /pmc/articles/PMC7292379/?report=abstract

5. Lu R, Wu X, Wan Z, Li Y, Jin X, Zhang C. A novel reverse transcription loop-mediated isothermal amplification method for rapid detection of sars-cov-2. Int J Mol Sci [Internet]. 2020 Apr 2 [cited 2020 Nov 27];21(8). Available from: /pmc/articles/PMC7216271/?report=abstract

6. Yan C, Cui J, Huang L, Du B, Chen L, Xue G, et al. Rapid and visual detection of 2019 novel coronavirus (SARS-CoV-2) by a reverse transcription loop-mediated isothermal amplification assay. Clin Microbiol Infect [Internet]. 2020 Jun 1 [cited 2020 Nov 27];26(6):773–9. Available from: /pmc/articles/PMC7144850/?report=abstract

7. Baek YH, Um J, Antigua KJC, Park JH, Kim Y, Oh S, et al. Development of a reverse transcription-loop-mediated isothermal amplification as a rapid early-detection method for novel SARS-CoV-2. Emerg Microbes Infect [Internet]. 2020 Jan 1 [cited 2020 Oct 11];9(1):998–1007. Available from: /pmc/articles/PMC7301696/?report=abstract

8. Chow FWN, Chan TTY, Tam AR, Zhao S, Yao W, Fung J, et al. A rapid, simple, inexpensive, and mobile colorimetric assay covid-19-lamp for mass on-site screening of covid-19. Int J Mol Sci [Internet]. 2020 Aug 1 [cited 2020 Oct 11];21(15):1–10. Available from: /pmc/articles/PMC7432162/?report=abstract

9. Huang WE, Lim B, Hsu C, Xiong D, Wu W, Yu Y, et al. RT-LAMP for rapid diagnosis of coronavirus SARS-CoV-2. Microb Biotechnol [Internet]. 2020 Jul 25 [cited 2020 Nov 26];13(4):950–61. Available from: https://onlinelibrary.wiley.com/doi/abs/10.1111/1751-7915.13586

10. Moher D, Liberati A, Tetzlaff J, Altman DG, Altman D, Antes G, et al. Preferred reporting items for systematic reviews and meta-analyses: The PRISMA statement. PLoS Med. 2009;6(7).

11. Reitsma JB, Leeflang MMG, Sterne JAC, Bossuyt PMM, Whiting PF, Rutjes AWSS, et al. Research and reporting methods accuracy studies. Ann Intern Med. 2011;155(4):529–36.

12. Rödel J, Egerer R, Suleyman A, Sommer-Schmid B, Baier M, Henke A, et al. Use of the variplex ™ SARS-CoV-2 RT-LAMP as a rapid molecular assay to complement RT-PCR for COVID-19 diagnosis. J Clin Virol [Internet]. 2020 Nov 1 [cited 2020 Oct 11];132:104616. Available from: /pmc/articles/PMC7457909/?report=abstract

13. Klein S, Müller TG, Khalid D, Sonntag-Buck V, Heuser AM, Glass B, et al. SARS-CoV-2 RNA extraction using magnetic beads for rapid large-scale testing by RT-qPCR and RT-LAMP. Viruses [Internet]. 2020 Aug 1 [cited 2020 Oct 11];12(8). Available from: /pmc/articles/PMC7472728/?report=abstract

14. Kitagawa Y, Orihara Y, Kawamura R, Imai K, Sakai J, Tarumoto N, et al. Evaluation of rapid diagnosis of novel coronavirus disease (COVID-19) using loop-mediated isothermal amplification. J Clin Virol [Internet]. 2020 Aug 1 [cited 2020 Oct 11];129:104446. Available from: /pmc/articles/PMC7241399/?report=abstract

15. Lau YL, Ismail I, Mustapa NI, Lai MY, Tuan Soh TS, Hassan A, et al. Real-time reverse transcription loop-mediated isothermal amplification for rapid detection of SARS-CoV-2. PeerJ [Internet]. 2020 Jun 3 [cited 2020 Nov 27];2020(6):e9278. Available from: https://peerj.com/articles/9278

16. Mohon AN, Oberding L, Hundt J, van Marle G, Pabbaraju K, Berenger BM, et al. Optimization and clinical validation of dual-target RT-LAMP for SARS-CoV-2. J Virol Methods [Internet]. 2020 Dec 1 [cited 2020 Nov 27];286:113972. Available from: /pmc/articles/PMC7490281/?report=abstract

17. Lee JYH, Best N, McAuley J, Porter JL, Seemann T, Schultz MB, et al. Validation of a single-step, single-tube reverse transcription loop-mediated isothermal amplification assay for rapid detection of SARS-CoV-2 RNA. J Med Microbiol [Internet]. 2020 Aug 5 [cited 2020 Oct 11];69(9). Available from: https://pubmed.ncbi.nlm.nih.gov/32755529/

18. Jiang M, Pan W, Arasthfer A, Fang W, Ling L, Fang H, et al. Development and Validation of a Rapid, Single-Step Reverse Transcriptase Loop-Mediated Isothermal Amplification (RT-LAMP) System Potentially to Be Used for Reliable and High-Throughput Screening of COVID-19. Front Cell Infect Microbiol [Internet]. 2020 Jun 16 [cited 2020 Oct 11];10:331. Available from: /pmc/articles/PMC7313420/?report=abstract

19. Yan C, Cui J, Huang L, Du B, Chen L, Xue G, et al. Rapid and visual detection of 2019 novel coronavirus (SARS-CoV-2) by a reverse transcription loop-mediated isothermal amplification assay. Clin Microbiol Infect [Internet]. 2020 Jun 1 [cited 2020 Oct 11];26(6):773–9. Available from: /pmc/articles/PMC7144850/?report=abstract

20. Subsoontorn P, Lohitnavy M, Kongkaew C. The diagnostic accuracy of nucleic acid point-of-care tests for human coronavirus: A systematic review and meta-analysis. Sci Rep [Internet]. 2020;1–13. Available from: https://doi.org/10.1038/s41598-020-79237-7

21. Sahoo PR, Sethy K, Mohapatra S, Panda D. Loop mediated isothermal amplification: An innovative gene amplification technique for animal diseases. Vet World [Internet]. 2016 May 11 [cited 2021 Jan 3];9(5):465–9. Available from: /pmc/articles/PMC4893716/?report=abstract

22. Ganguli A, Mostafa A, Berger J, Aydin MY, Sun F, Stewart de Ramirez SA, et al. Rapid isothermal amplification and portable detection system for SARS-CoV-2. Proc Natl Acad Sci U S A [Internet]. 2020 Sep 15 [cited 2020 Oct 11];117(37):22727–35. Available from: /pmc/articles/PMC7502724/?report=abstract

23. Farooq U, Latif ;, Irshad ;, Ullah ;, Zahur ;, Naeem ;, et al. Loop-mediated isothermal amplification (RT-LAMP): a new approach for the detection of foot-and-mouth disease virus and its sero-types in Pakistan. Vol. 16. 2015.

24. Kim JH, Kang M, Park E, Chung DR, Kim J, Hwang ES. A Simple and Multiplex Loop-Mediated Isothermal Amplification (LAMP) Assay for Rapid Detection of SARS-CoV. Biochip J [Internet]. 2019 Dec 1 [cited 2021 Jan 4];13(4):341–51. Available from: /pmc/articles/PMC7097549/?report=abstract

25. Huang P, Wang H, Cao Z, Jin H, Chi H, Zhao J, et al. A rapid and specific assay for the detection of MERS-CoV. Front Microbiol [Internet]. 2018 May 29 [cited 2021 Jan 4];9(MAY). Available from: /pmc/articles/PMC5987675/?report=abstract

26. Thompson D, Lei Y. Mini review: Recent progress in RT-LAMP enabled COVID-19 detection. Sensors and Actuators Reports [Internet]. 2020;2(1):100017. Available from: https://doi.org/10.1016/j.snr.2020.100017

27. Kashir J, Yaqinuddin A. Loop mediated isothermal amplification (LAMP) assays as a rapid diagnostic for COVID-19. Med Hypotheses [Internet]. 2020 Aug 1 [cited 2020 Oct 11];141:109786. Available from: /pmc/articles/PMC7182526/?report=abstract

